# Likely cross-border introductions of MPXV Clade I into the Republic of the Congo from the Democratic Republic of the Congo

**DOI:** 10.1101/2024.08.21.24312265

**Authors:** Claude Kwe Yinda, Félix Koukouikila-Koussounda, Pembe Issamou Mayengue, Golmard Reiche Elenga, Benjamin Greene, Missiani Ochwoto, Ghislain Dzeret Indolo, Yanne Vanessa Thiécesse Mavoungou, Dachel Aymard Eyenet Boussam, Bani Reize Vishnou Ampiri, Chastel Claujens Mapanguy Mfoutou, Yvanhe Deho Kianguebeni Mbouala, Francine Ntoumi, Jean-Médard Kankou, Vincent J. Munster, Fabien Roch Niama

## Abstract

**Background:** Mpox virus (MPXV) Clade I is endemic in several central African countries and characterized by severe disease and high mortality. Since October 2023, a large-scale Mpox outbreak has emerged in the Democratic Republic of the Congo (DRC), with 22,477 cases and 1,038 deaths reported as of May 26, 2024, and World Health organization (WHO) has declared mpox a public health emergency of international concern on august 14 2024. Twenty-three provinces in the DRC have been affected, 15 of which border another country, including the Republic of the Congo (RoC). The RoC is bordered by five of these affected provinces, increasing the risk of cross-border spread. In the RoC, Mpox cases have historically occurred in the endemic areas of the Likouala department in the north. Recently, suspected cases have emerged outside this region, and it is unclear if these cases are recent spillover events from the DRC outbreaks. Therefore, we performed molecular analyses of these cases in the RoC.

**Methods:** Samples were collected from suspected cases between January and April 29, 2024, in Likouala, Cuvette-Centralle, and Pointe-Noire. Blood samples, skin/oropharyngeal swabs or skin crusts were obtained for molecular diagnosis at the Laboratoire National de Santé Publique (LNSP), Brazzaville. MPXV sequences were obtained and analyzed using newly established Nanopore sequencing methodology and bioinformatic pipeline.

**Findings:** Phylogenetic analysis of sequences shows the clustering of MPXV sequences obtained from cases in RoC with sequences from the ongoing Mpox outbreak in the DRC. In addition, sequences from the RoC show multiple phylogenetic positioning suggesting the occurrence of multiple co-circulating strains in the human population.

**Interpretation:** The close genetic relatedness between sequences from RoC and those from DRC indicates a possible cross-border transmission of MPXV from DRC to RoC. These data highlight the need for implementation of expanded surveillance in countries bordering DRC and RoC, in combination with control measures focused at containing the current outbreaks in DRC and RoC to prevent escalation into a larger-scale epidemic.

**Funding:** This research was supported by the Intramural Research Program of the National Institute of Allergy and Infectious Diseases (NIAID), National Institutes of Health (NIH).

## Introduction

Mpox virus (MPXV) formerly known as monkeypox virus, is an enveloped, double stranded DNA virus of the *Poxviridae* family and the etiological agent of Mpox ^1^. There are two known clades of MPXV: clade I which is endemic in central Africa and has been detected in the Democratic Republic of the Congo (DRC), the Republic of the Congo (RoC), the Central African Republic, Cameroon and Gabon, and clade II which is endemic in West Africa ^2^. Mpox typically presents with a 2–4-day prodrome followed by the appearance of a vesiculo-papular rash and is combined with fever and lymphadenopathy ^3^. Case fatality rates (CFRs) across previously reported Mpox outbreaks have averaged 11% for clade I MPXV and <4 % for clade II MPXV ^4,5^. In May 2022, Clade IIb MPXV caused the largest known multinational human outbreak of Mpox with over 92,590 laboratory-confirmed cases, mostly outside of historically endemic regions in West Africa ^6^. The Clade IIb MPXV outbreak appears to be largely driven by sustained human-to-human transmission via male-to-male sexual or intimate contact (MSM). This Mpox outbreak in non-endemic countries resulted in the declaration of a public health emergency of international concern by the World Health Organization (WHO) in 2022 ^6^.

The RoC, a country of 342,000 km^2^ with an estimated population size of 5.7 million inhabitants, is endemic for Clade I MPXV in the northern part of the country ^7,8^, and since the first confirmed cases reported in 2003 ^9^ small self-limiting outbreaks have been reported over the last couple of years. This historic occurrence of small outbreaks in RoC is in contrast with the continuous increase in cases over the last decades, and the surge of cases throughout most of DRC reported over 2023 and 2024 ^10-13^. Genomic analysis revealed a novel distinct MPXV clade now designated as Clade Ib, divergent from previously sequenced Clade I strains circulating in eastern DRC ^10^. The recent increase in Clade I Mpox cases suggest an increase in human-to-human transmissibility for Clade I, similar to what has been reported for Clade IIb. There are clear similarities between the emergence of Clade IIb and Clade I, with sexual contact transmission now being reported for Clade I in DRC ^14^. However, whereas the clade IIb MPXV circulated almost exclusively within the MSM population, the Clade I MPXV displays broad circulation throughout the human population ^15 14^. Due to this upsurge of Mpox in the DRC and cases outside Africa ^16^, the WHO on August 14, 2024 declared Mpox a public health emergency of international concern for the second time ^17^.

In March 2024, the first Mpox cases were reported outside the historic endemic areas in the RoC and it is not clear if these cases are recent spillover events from the DRC outbreaks. Here we describe the sequencing analyses of samples obtained from the Mpox cases in RoC at the Laboratoire National de Santé Publique (LNSP), Brazzaville.

## Methods

### Study design

We performed a cross-sectional descriptive study of genomic sequences analyses on MPXV real-time PCR confirmed samples collected in different sanitary departments in the RoC from January to April 2024. MPXV positive samples were consecutively selected regardless of the cycle threshold (Ct) values. Further selection criteria included the quantity of the remaining biological sample after real-PCR screening and the availability of the filled data collection form. The study took place at the molecular biology unit of the LNSP, Brazzaville.

### Sampling and data collection

Mpox is one of the 18 diseases under surveillance in the RoC. In 2022, the Ministry of Health and Population set a passive surveillance program for Mpox across the Country. Under this program, focal points were trained for recognition of suspected cases following criteria for Mpox suspicious set by the Direction de l’Epidémiologie et de la Lutte contre la Maladie (DELM), sampling, data collection using a national standardized form established by the DELM and sample shipment to the LNSP which is the national reference laboratory. If a case is confirmed, a team including biologists from LNSP and epidemiologists from DELM is commissioned to go into the outbreak area to conduct intensive field investigations consisting of identification of contacts and that of new suspected cases, data collection, sampling (including whole blood, skins lesion crusts or skin swabs) and rapid samples’ transfer to the LNSP for molecular diagnosis.

From January to April 2024, a total of 61 samples collected from suspected cases were received at the LNSP. Suspected cases were from Brazzaville (9 cases), Cuvette centrale (23 cases), Likouala (10 cases), Plateaux (4 cases) and Pointe-Noire (25 cases).

### Real-time PCR screening of the samples

For all the samples, samples were handled in a class III biosafety cabinet, DNA was extracted using the QIAamp DNA Mini Kit (Qiagen, Hilden, Germany), following the manufacturer’s instructions. Extracted DNA was subsequently used for molecular diagnosis. Real time PCR assays were performed using the Monkeypox Virus Real Time PCR Kit (Shanghai ZJ Bio-Tech Co., Ltd. (“Liferiver”), China) according to manufacturer’s instructions. Of the 61 samples, MPXV DNA was detected in 35 (57.37%), and based on the study criteria, 31 samples were selected and stored at −20°C until further used for genomic sequencing under the current study.

### MPXV sequencing

#### Amplicon sequencing

For sequencing of the positive samples, we used previously published methods (Ochwoto et al., 2024, in preparation). Briefly, after the presence of PCR products were verified using gel electrophoresis, the five PCR products were pooled together, and subject to end repair (NEB, US) and barcoding (ONT, UK). Next, samples were cleaned with 0.4x volume AMPure XP beads (Beckman, US) and eluted in 30 µL of 10 millimolar Tris pH 8.0. Then sequencing adapters were ligated to the barcoded amplicons using 5 µL of Adapter Mix AMII (ONT), 5 µL of Quick T4 DNA Ligase (NEB, US) and 10 µL of NEBNext Quick Ligation Reaction Buffer 5x (NEB, US) to 30 µL of the barcoded amplicon pool. Following a twenty-minute incubation at room temperature, samples were cleaned again using AMPure XP beads (Beckman) and DNA was eluted in 15 µL of Elution Buffer (ONT, UK). DNA libraries were then quantified using the Qubit High Sensitivity dsDNA assay according to manufacturer’s instructions (Thermofisher, US). Libraries were sequenced on Oxford Nanopore’s GridION (ONT, UK) using FLO-MIN106-D flow cell (ONT, UK) in accordance with the manufacturer’s protocol. FASTQ files were processed through Mpox-seek pipeline (https://openomics.github.io/Mpox-seek/#citation) for read quality control, adapter trimming, consensus sequence, and phylogenetic tree generation.

#### Full genome sequencing

To determine the type of single nucleotide polymorphisms (SNPs) in these novel sequences we further recovered the complete genomes of the sequences. First, the quality of the DNA was assessed on the Agilent Tapestation 4200 (Agilent Technologies, Inc., USA) using genomic tapes/reagents. One hundred nanograms of DNA was sheared to an average size of 550bp using the Covaris LE220 (Covaris, LLC, MA) and final libraries were prepared using the Illumina TruSeq DNA Nano v2 protocol (Illumina, USA) with 8 cycles of PCR amplification using 6ul of IDT/Illumina-TruSeq UD Indexes as barcoded primers. Final libraries were cleaned using an 80% vol of AMPureXP beads (Agencourt, USA), visualized on a BioAnalyzer HS chip (Agilent Technologies, Inc., USA), and quantified using the Kapa SYBR FAST Universal qPCR kit for Illumina sequencing (Kapa Biosystems, USA) on the CFX384 Real-Time PCR Detection System (Bio-Rad Laboratories, Inc, USA) and Qubit Fluorometer (Thermo Fisher Scientific, USA). Libraries consisting of 40-250 ng of DNA were pooled into three pools (according to amount of DNA and Ct value of sample). Viral sequences were captured using the HyperCap enrichment kit (Roche Diagnostic Corp., USA), and final library was cleaned, analyzed using the Agilent bioanalyzer and quantified using the Kapa SYBR FAST Universal qPCR as above. Each of the 3 captured-library pools were diluted to a final concentration of 2 nM and pooled equimolarly. This final, MPXV DNA-captured, library pool was denatured and diluted to a 10.5 pM stock and a paired-end 2 x 250 cycle sequencing was carried out on the MiSeq using a V2 flow cell and 500 cycle chemistry.

After sequencing, raw fastq reads were trimmed of adapter sequences using Cutadapt v. 1.12 ^18^ and filtered for quality using Fastx Toolkit v. 0.0.14 ^19^. Reads were then screened against the human genome to remove host reads. Remaining reads for one of the sample were then de novo assembled using SPAdes v. 3.15.5 ^20^. Resulting contigs were manually assembled to create the full-length genome. Assembly errors were corrected automatically using multiple rounds of pilon v. 1.22 ^21^ and manual correction. The resulting assembly was then used for reference assembly of the remaining genomes using Bowtie2 v. 2.2.9 ^22^ followed by multiple rounds of correction using pilon v. 1.22.

#### Phylogenetic analysis

Full length genomes of old Clade I MPXV sequences were obtained from GISAID database (https://www.gisaid.org/) and recent sequences from the Kamituga outbreak were obtained from Github (https://github.com/inrblabgenpath/Mpox_sequencing_Kamituga). Clade IIb MPXV from GISAID were used as outgroup. All sequences were trimmed to the required region and alignments were built using MAFFT (FFT-NS-1 algorithm) ^23^, with the best model for distance estimates identified with the ModelFinder function ^24^ as the one with the lowest Bayesian information criterion (BIC). Maximum likelihood phylogenetic tree was constructed using IG-TREE2 ^25^ and branch support was assessed using both ultrafast bootstrap approximation (ufBoot, 1000 replicates) ^26^ and SH-like approximate likelihood ratio test (SH-aLRT). The tree was visualized in FigTree (http://tree.bio.ed.ac.uk/software/figtree/) and rooted using Clade IIb sequences. Only bootstrap values greater than 69% are shown. Bars indicate nucleotide substitutions per site.

Furthermore, we utilized the full genomes to perform ancestral reconstruction for each internal node on phylogeny, enabling mapping of single nucleotide polymorphisms (SNPs) along branches following recently developed method ^27^. SNPs were categorized according to their consistency with the APOBEC3 editing signature, which is characterized by specific mutations (TC → TT and GA → AA). To estimate the date when the non-Kamituga MPXV genomes (putative Clade Ib) outbreak in DRC and RoC started, we excluded all Kamituga MPXV genomes and assumed non-APOBEC3 rate of evolution and used strict molecular clock with constant model implemented in BEAST v. 1.10.4 ^28^. For each analysis, three independent chains of 100 million generations (sampled every 100,000 states) were run to ensure convergence and then combined with appropriate burn-in. Statistical uncertainty was reflected in values of the 95% highest probability density (HPD). The chains were then combined with LogCombiner v. 1.10.4. For all subsequent analyses, we assessed convergence using Tracer, and constructed a maximum clade credibility (MCC) tree (estimated from the posterior distribution of trees with node heights scaled to mean values and posterior probabilities showing the statistical support for individual nodes) in TreeAnnotator v. 1.10.4.

## Results

A total of 31 Mpox confirmed cases between January and April 29, 2024, were included in this study. Demographic data of these participants are summarized in Table 1. Mpox positive cases were from Cuvette Centrale (19/31, 61%), Likouala (8/31, 26%), and Pointe Noire (5/31, 16%) (Figure 1A) with males being the majority of these confirmed MPXV infection cases (20/31, 65%). The median age was 20 years (IQR 11-31) (Table 1). Children under 15 years constituted 39% (12/31) of confirmed cases, individuals aged 15-30 years made up 35% (11/31), and those above 30 years accounted for 23% (7/31) of the cases. None of the individuals with confirmed Mpox had been vaccinated against smallpox.

**Table 1:** Demographic characteristics of Mpox confirmed cases.

| ID | Sequence ID/Accessions | Date | Gender | Age range | Department | Sample type | Ct-value | Full genome recovery |
| --- | --- | --- | --- | --- | --- | --- | --- | --- |
| MPOX-CG-006 | hMpxV/ROC/Cuvette-Centrale/2024/01 | Feb-24 | M | 11-15 | Cuvette-Centrale | Skin lesion | 16.43 | Y |
| MPOX-CG-010 | hMpxV/ROC/Cuvette-Centrale/2024/02 | Mar-24 | M | 6-10 | Cuvette-Centrale | Skin lesion | 21.78 | Y |
| MPOX-CG-014 | hMpxV/ROC/Cuvette-Centrale/2024/03 | Feb-24 | F | 36 - 40 | Cuvette-Centrale | Skin lesion | 18.26 | Y |
| MPOX-CG-015 | hMpxV/ROC/Cuvette-Centrale/2024/04 | Mar-24 | M | 26-30 | Cuvette-Centrale | Skin lesion | 16.43 | Y |
| MPOX-CG-023 | hMpxV/ROC/Cuvette-Centrale/2024/05 | Mar-24 | F | 0-5 | Cuvette-Centrale | Skin lesion | 20.29 | Y |
| MPOX-CG-025 | hMpxV/ROC/Cuvette-Centrale/2024/06 | Mar-24 | F | 21-25 | Cuvette-Centrale | Skin lesion | 15.96 | Y |
| MPOX-CG-043 | hMpxV/ROC/Likouala/2024/07 | Feb-24 | F | 6-10 | Likouala | Skin crust | 15.42 | Y |
| MPOX-CG-045 | hMpxV/ROC/Likouala/2024/08 | Feb-24 | M | NR | Likouala | Skin crust | 16.82 | Y |
| MPOX-CG-061 | hMpxV/ROC/Likouala/2024/09 | Mar-24 | M | 36-40 | Likouala | Skin crust | 13.36 | Y |
| MPOX-CG-062 | hMpxV/ROC/Likouala/2024/010 | Mar-24 | M | 36-40 | Likouala | Skin lesion | 19.01 | Y |
| MPOX-CG-011 | hMpxV/ROC/Cuvette-Centrale/2024/011 | Feb-24 | F | 26-30 | Cuvette-Centrale | Skin lesion | 34.34 | N |
| MPOX-CG-013 | hMpxV/ROC/Cuvette-Centrale/2024/012 | Feb-24 | M | 6-10 | Cuvette-Centrale | Skin lesion | 26.46 | Y |
| MPOX-CG-017 | hMpxV/ROC/Cuvette-Centrale/2024/013 | Mar-24 | M | 46-50 | Cuvette-Centrale | Skin lesion | 22.78 | Y |
| MPOX-CG-026 | hMpxV/ROC/Cuvette-Centrale/2024/014 | Mar-24 | F | 21-25 | Cuvette-Centrale | Blood | 33.74 | N |
| MPOX-CG-027 | hMpxV/ROC/Cuvette-Centrale/2024/015 | Feb-24 | F | 36-40 | Cuvette-Centrale | Blood | 33.74 | N |
| MPOX-CG-035 | hMpxV/ROC/Likouala/2024/16 | Feb-24 | M | 11-15 | Likouala | Skin lesion | 32.39 |  |
| MPOX-CG-036 | hMpxV/ROC/Likouala/2024/017 | Feb-24 | M | NR | Likouala | Skin lesion | 21.78 | Y |
| MPOX-CG-042 | hMpxV/ROC/Likouala/2024/018 | Feb-24 | F | 6-10 | Likouala | Skin crust | 23.29 | Y |
| MPOX-CG-044 | hMpxV/ROC/Likouala/2024/019 | Feb-24 | M | 11-15 | Likouala | Skin crust | 27.2 | N |
| MPOX-CG-046 | hMpxV/ROC/Pointe-Noire/2024/020 | Feb-24 | M | 16-20 | Pointe-Noire | Blood | 33.44 | N |
| MPOX-CG-019 | NS | Mar-24 | F | 26-30 | Cuvette-Centrale | Skin lesion | 37.44 | N |
| MPOX-CG-020 | NS | Mar-24 | F | 26-30 | Cuvette-Centrale | Blood | 35.96 | N |
| MPOX-CG-022 | NS | Mar-24 | M | 26-30 | Cuvette-Centrale | Blood | 35.83 | N |
| MPOX-CG-048 | NS | Feb-24 | M | 16-20 | Pointe-Noire | Skin lesion | 34.71 | N |
| MPOX-CG-049 | NS | Feb-24 | F | 11-15 | Pointe-Noire | Skin crust | 35.56 | N |
| MPOX-CG-024 | NS | Feb-24 | F | 0-5 | Cuvette-Centrale | Blood | 34.34 | N |
| MPOX-CG-007 | NS | Mar-24 | M | 11-15 | Cuvette-Centrale | Blood | 36 | N |
| MPOX-CG-012 | NS | Feb-24 | M | 0-5 | Pointe-Noire | Skin lesion | 34.34 | N |
| MPOX-CG-008 | NS | Feb-24 | M | 31-35 | Cuvette-Centrale | Blood | 37.09 | N |
| MPOX-CG-016 | NS | Feb-24 | M | 26-30 | Cuvette-Centrale | Blood | 35.96 | N |
| MPOX-CG-018 | NS | Mar-24 | M | 46-50 | Cuvette-Centrale | Blood | 35.83 | N |

NR: age range not recorded, NS: No sequences obtained due to low viral load or poor sequence quality, M = male, F = female, Y = full genome recovery successful, N = full genome recovery failed.

**Figure 1:**
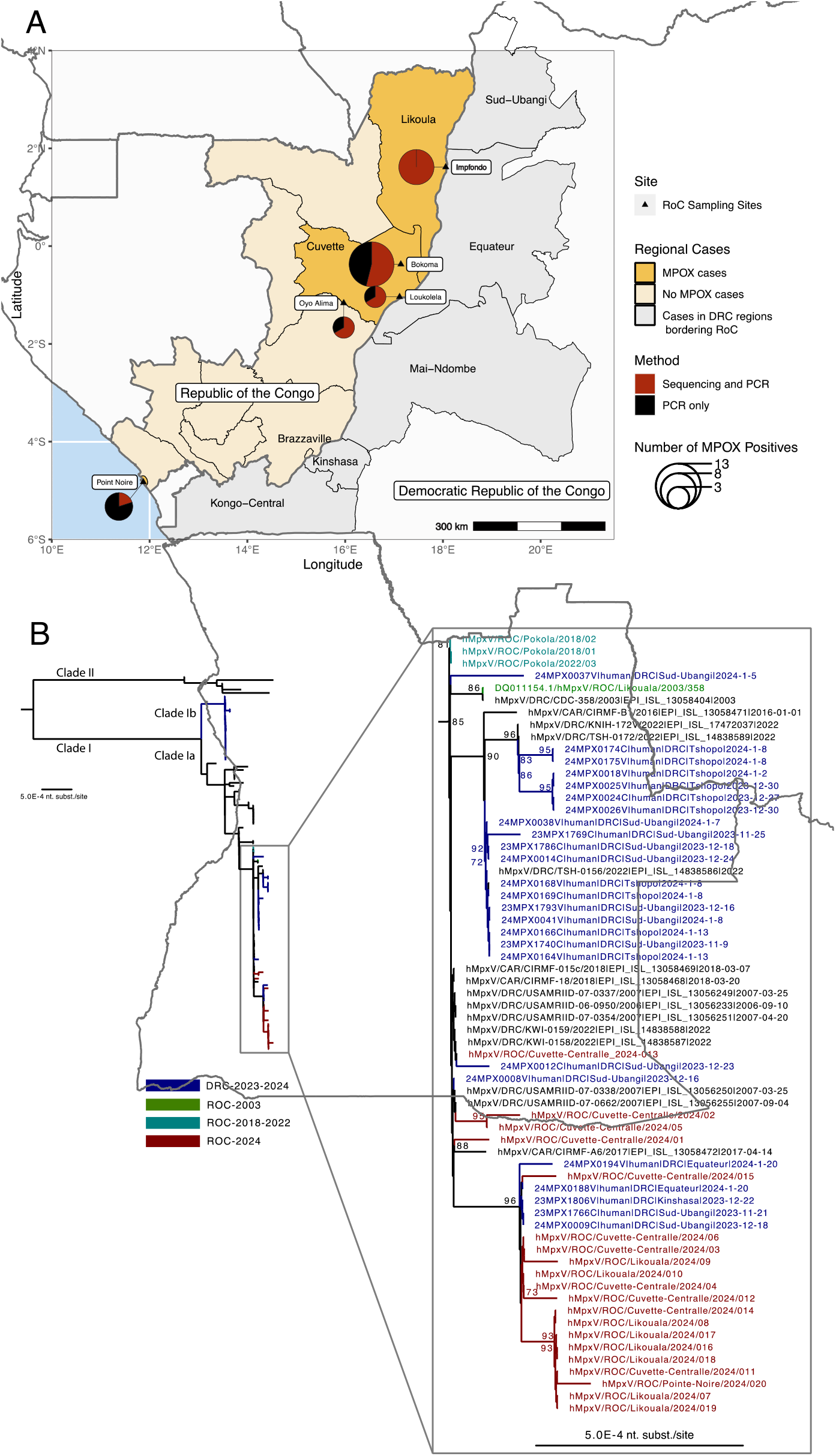
**A**. Map of the Republic of the Congo (RoC) and the neighboring provinces of the Democratic Republic of the Congo (DRC). RoC regions with Mpox positive cases in yellow and DRC grey. Size of the circle correspond with the number of cases, whereas the red to black ratio indicates the number of Mpox PCR positive samples for which sequencing information was obtained. **B**. Phylogenetic analysis of MPXV nucleotide sequences (∼12kb from the 3’ end) including historic and contemporary Clade I sequence. Alignments were built with MAFFT (FFT-NS-1 algorithm) ^23^, with the best model for distance estimates identified with the ModelFinder function ^24^ as the one with the lowest Bayesian information criterion (BIC). Maximum likelihood phylogenetic tree was constructed using IG-TREE2 ^25^ and branch support was assessed using both ultrafast bootstrap approximation (ufBoot, 1000 replicates) ^26^ and SH-like approximate likelihood ratio test (SH-aLRT). The tree was visualized in FigTree (http://tree.bio.ed.ac.uk/software/figtree/) and the phylogenetic tree were rooted using Clade II MPXV sequences. MPXV sequences obtained in this study are indicated in red. Whereas, historic RoC sequences are indicated in green, and contemporary DRC sequences from the current outbreaks are indicated in blue ^30^. Node labels represent bootstrap values greater than 69, 1,000 replicates. Scale bar indicates nucleotide substitutions per site.

Overall, cycle threshold (Ct) ranged between 13.4 and 37.4. Respectively, we recovered eleven, eight and one sequence from positive samples from Cuvette Centrale, Likouala and Pointe Noire Provinces (Figure 1A). Phylogenetic analysis (Figure 1B) revealed that MPXV strains sequenced from RoC are found in two major clusters within Clade Ia. The first is a more diverse cluster made up of four sequences from this study all recovered from samples from the RoC Cuvette Province clustering with two MPXV sequences from the current DRC outbreak (in Sud-Ubangi), three older sequences from Central African Republic sequenced between 2017 and 2018, and seven sequences from DRC sequenced in 2006-07 and 2022. The second cluster is made up of 16 sequences from this study clustering with sequences from the current DRC outbreak (two sequences from Sud-Ubangil, two from Equateur and one from Kinshasa). The nucleotide similarity between sequences from RoC and DRC in this cluster ranged from 99.5 to 99.9%. Sequences from 2018-2022 MPXV samples from the Pokola in the Sangha Province in RoC clustered together with each other.

To analyze single nucleotide polymorphisms (SNPs) in these new sequences from the RoC, we subjected the MPXV DNA from these samples to full genome sequencing, which produced 14 complete genomes: 8 from Cuvette Province and 6 from Likouala Province. All 14 genomes displayed consistent phylogenetic placement with the ∼13Kb sequences (Supplementary Figure 1A). Additionally, the proportion of APOBEC3-type mutations relative to other mutations in these sequences was 16.2% (11 of 68 observed mutations) (Supplementary Figure 1B). To estimate when these recent sequences from RoC were introduced into the human population, we conducted molecular clock analysis. This analysis revealed two clusters of sequences from RoC and the Democratic Republic of the Congo (DRC): the first cluster, which emerged around 2000, includes RoC sequences from Sangha (a past outbreak) and Cuvette (the current outbreak) regions, along with one DRC sequence from Sud-Ubangil Province. The second cluster, which emerged in 2005, comprises the remaining RoC samples from the current outbreak and five sequences from one of the ongoing DRC outbreaks, consistent with the ∼13Kb sequences (Supplementary Figure 2).

## Discussion

Human infection with MPXV was first recognized in 1970 in DRC, with historically sporadic Mpox outbreaks. The last decade has seen increasing frequency of Clade I Mpox cases in DRC ^11^. The rise in the number of Mpox cases in DRC in 2023, their occurrence in new geographic areas, including outside Africa ^10,29,30^ resulted in second time Mpox was declared Mpox a public health emergency of international concern by the WHO on August 14, 2024 ^17^. This increase in cases suggests either anthropogenic changes, behavior changes, virus changes, or a combination thereof driving the increased human-to-human transmission. During the ongoing Mpox epidemic in DRC, the neighboring RoC observed a sudden rise in the number of Mpox cases in 2024. This resulted to the declaration of a Mpox epidemic by the Ministry of Public Health of RoC in April 24, 2024 ^31^. Molecular epidemiology, using pathogen sequencing data has been successfully used in Africa to trace the spread of emerging zoonotic pathogens such as Ebola virus and Lassa virus ^32,33^. To understand whether the increase in Mpox cases in RoC was driven by local emergence of MPXV or spillover from DRC, we implemented a novel next-generation sequencing approach using the Oxford Nanopore GridION platform.

The sequences of the Clade Ia MPXV from the RoC, obtained during the current outbreak cluster roughly in three separate lineages. The diverse phylogenetic positioning of the RoC MPXV sequences suggests the occurrence of multiple co-circulating strains in the human population and reflects the MPXV diversity observed during the ongoing outbreaks in DRC ^10^. Genomic data from DRC demonstrates the co-circulation of multiple independent lineages, including Clade Ib, suggesting several independent spillover events from the natural reservoir ^30^. The phylogenetic congruence of circulating MPXV lineages observed in cases from the regions of the DRC adjacent to the RoC suggests multiple independent introductions of MPXV into human populations within the RoC. These introductions could have resulted from direct zoonotic transmission of similar viral strains from a reservoir host into populations in both the DRC and RoC or through cross-border human-to-human transmission. The latter is supported by the observed 16% frequency of APOBEC3-like mutations in these new sequences, a significant increase from the 8% (38 out of 463 sequences) previously reported in other Clade I sequences from the DRC and other endemic regions of Africa ^27^. This increase in APOBEC3-like mutations suggests a marginally higher rate of human-to-human transmission in these recent cases compared to earlier reports. Consequently, this finding strengthens the argument for cross-border human-to-human transmission as a significant factor in the spread of MPXV. Given the magnitude of the outbreaks in the DRC, it is highly likely that the cases in the RoC represent spillover events from the DRC into the RoC. However, the available epidemiological data is insufficient to definitively confirm the directionality of Mpox transmission. The border areas between RoC and DRC are known for extensive connectivity of the populations, with major waterways connecting the region and major population centers such Mbandaka, Kinshasa and Brazzaville and Lingala as the common language. Given the frequent travel between RoC and DRC, there exists an elevated risk of trans-border transmission. Moreover, the clustering of sequences from Mpox cases across different provinces in RoC, including Cuvette, Likouala, and Pointe Noire, suggests probable local human-to-human transmission during the ongoing outbreak in RoC, however, the rate of this transmission compared to the observed Clade IIb and Ib MPXV ^13,27^ might be lower.

The reported 35 cases likely represent an underestimate of the true prevalence of Mpox cases in RoC. Discussions with local healthcare workers reveal that many individuals exhibiting signs and symptoms of Mpox remain within the community without seeking medical attention. The drivers of the current human-to-human transmission in RoC are unknown, and follow-up epidemiological investigations and expanded surveillance are urgently needed to better understand the modes of MPXV transmission. In addition, it is currently unclear whether sexual contact transmission is contributing to the outbreak as has been observed in Sud Kivu region in DRC ^14^. The sustained human-to-human transmission of clade I MPXV in the DRC and the subsequent introduction into RoC, presents a significant cause for concern. This introduction into RoC from DRC, highlights the vulnerability of the region for cross border introductions potentially driving outbreaks over large geographical areas into bordering countries. We strongly advocate for increased international support to strengthen epidemiology and surveillance, real-time molecular tracking of MPXV in combination with broad scale availability of prophylactic and therapeutic countermeasures to prevent escalation into another global Mpox outbreak ^34^.

## Data Availability

All novel sequences reported are submitted to GISAID.

## Acknowledgments

This research was supported by the Intramural Research Program of the National Institute of Allergy and Infectious Diseases (NIAID), National Institutes of Health (NIH). We thank NIAID’s Research Technology Branch for whole genome sequencing (Craig Martens and Kent D. Barbian) and for developing bioinformatic pipeline (Skyler Kuhn and Paul Schaughency). We equally thank Dr. Willibrord “Chief Willy” Shasha (USAID) and Dr. Kerton R. Victory (CDC), for in country technical support. We are thankful to WHO Congo for supporting biological sample transport system and providing real-time PCR kits. We also appreciate the logistic support from the US Embassy in RoC.

## Ethics

This study was carried out in accordance with the declaration of Helsinki and was approved by the Congolese Foundation for Medical Research Ethics committee (Avis n°053/CEI/FCRM/2024).

## References

1. Hughes AL, Irausquin S, Friedman R. The evolutionary biology of poxviruses. Infection, Genetics and Evolution 2010; 10(1): 50–9.

2. McCollum AM, Shelus V, Hill A, et al. Epidemiology of Human Mpox - Worldwide, 2018-2021. MMWR Morb Mortal Wkly Rep 2023; 72(3): 68–72.

3. Altindis M, Puca E, Shapo L. Diagnosis of monkeypox virus - An overview. Travel Med Infect Dis 2022; 50: 102459.

4. Bunge EM, Hoet B, Chen L, et al. The changing epidemiology of human monkeypox-A potential threat? A systematic review. PLoS Negl Trop Dis 2022; 16(2): e0010141.

5. Hens M, Brosius I, Berens-Riha N, et al. Characteristics of confirmed mpox cases among clinical suspects: A prospective single-centre study in Belgium during the 2022 outbreak. New Microbes New Infect 2023; 52: 101093.

6. WHO. WHO Director-General declares the ongoing monkeypox outbreak a Public Health Emergency of International Concern. 2022. https://www.who.int/europe/news/item/23-07-2022-who-director-general-declares-the-ongoing-monkeypox-outbreak-a-public-health-event-of-international-concern (accessed 9/19/2022 2022).

7. Doshi RH, Guagliardo SAJ, Dzabatou-Babeaux A, et al. Strengthening of Surveillance during Monkeypox Outbreak, Republic of the Congo, 2017. Emerg Infect Dis 2018; 24(6): 1158–60.

8. Doshi RH, Guagliardo SAJ, Doty JB, et al. Epidemiologic and Ecologic Investigations of Monkeypox, Likouala Department, Republic of the Congo, 2017. Emerg Infect Dis 2019; 25(2): 281–9.

9. Learned LA, Reynolds MG, Wassa DW, et al. Extended interhuman transmission of monkeypox in a hospital community in the Republic of the Congo, 2003. Am J Trop Med Hyg 2005; 73(2): 428–34.

10. Masirika LM, Udahemuka JC, Schuele L, et al. Ongoing mpox outbreak in Kamituga, South Kivu province, associated with monkeypox virus of a novel Clade I sub-lineage, Democratic Republic of the Congo, 2024. Euro Surveill 2024; 29(11).

11. Ladnyj ID, Ziegler P, Kima E. A human infection caused by monkeypox virus in Basankusu Territory, Democratic Republic of the Congo. Bull World Health Organ 1972; 46(5): 593–7.

12. Rimoin AW, Mulembakani PM, Johnston SC, et al. Major increase in human monkeypox incidence 30 years after smallpox vaccination campaigns cease in the Democratic Republic of Congo. Proceedings of the National Academy of Sciences 2010; 107(37): 16262–7.

13. Vakaniaki EH, Kacita C, Kinganda-Lusamaki E, et al. Sustained human outbreak of a new MPXV clade I lineage in eastern Democratic Republic of the Congo. Nature Medicine 2024.

14. Kibungu E, Vakaniaki E, Kinganda-Lusamaki E, et al. Clade I–Associated Mpox Cases Associated with Sexual Contact, the Democratic Republic of the Congo. Emerging Infectious Disease journal 2024; 30(1): 172.

15. Web R. DR Congo: Rapport de la situation épidémiologiue de la variole simienne - sitrep no 2, 5-11 février 2024, 2024.

16. Control) EECfDPa. Risk assessment for the EU/EEA of the mpox epidemic caused by monkeypox virus clade I in affected African countries. 2024.

17. Organization WH. WHO Director-General declares mpox outbreak a public health emergency of international concern. 2024. p. https://www.who.int/news/item/14-08-2024-who-director-general-declares-mpox-outbreak-a-public-health-emergency-of-international-concern.

18. Martin M. Cutadapt removes adapter sequences from high-throughput sequencing reads. 2011 2011; 17(1): 3.

19. Gordon A. FASTX-Toolkit (RRID:SCR_005534). http://hannonlab.cshl.edu/fastx_toolkit/.

20. Bankevich A, Nurk S, Antipov D, et al. SPAdes: a new genome assembly algorithm and its applications to single-cell sequencing. J Comput Biol 2012; 19(5): 455–77.

21. Walker BJ, Abeel T, Shea T, et al. Pilon: An Integrated Tool for Comprehensive Microbial Variant Detection and Genome Assembly Improvement. PLOS ONE 2014; 9(11): e112963.

22. Langmead B, Salzberg SL. Fast gapped-read alignment with Bowtie 2. Nat Methods 2012; 9(4): 357–9.

23. Katoh K, Standley DM. MAFFT multiple sequence alignment software version 7: improvements in performance and usability. Mol Biol Evol 2013; 30(4): 772–80.

24. Kalyaanamoorthy S, Minh BQ, Wong TKF, von Haeseler A, Jermiin LS. ModelFinder: fast model selection for accurate phylogenetic estimates. Nat Methods 2017; 14(6): 587–9.

25. Minh BQ, Schmidt HA, Chernomor O, et al. IQ-TREE 2: New Models and Efficient Methods for Phylogenetic Inference in the Genomic Era. Molecular Biology and Evolution 2020; 37(5): 1530–4.

26. Hoang DT, Chernomor O, von Haeseler A, Minh BQ, Vinh LS. UFBoot2: Improving the Ultrafast Bootstrap Approximation. Mol Biol Evol 2018; 35(2): 518–22.

27. O’Toole Á, Neher RA, Ndodo N, et al. APOBEC3 deaminase editing in mpox virus as evidence for sustained human transmission since at least 2016. Science 2023; 382(6670): 595–600.

28. Drummond AJ, Rambaut A. BEAST: Bayesian evolutionary analysis by sampling trees. BMC Evol Biol 2007; 7: 214.

29. Organization WH. Multi-country outbreak of mpox. External situation report#30 – 25 November 2023., 2023.

30. Vakaniaki EH, Kacita C, Kinganda-Lusamaki E, et al. Sustained Human Outbreak of a New MPXV Clade I Lineage in Eastern Democratic Republic of the Congo. medRxiv 2024: 2024.04.12.24305195.

31. Reuters. Congo Republic declares mpox epidemic. 2024. p. ROC mpox outbreak.

32. Arias A, Watson SJ, Asogun D, et al. Rapid outbreak sequencing of Ebola virus in Sierra Leone identifies transmission chains linked to sporadic cases. Virus Evol 2016; 2(1): vew016.

33. Kafetzopoulou LE, Pullan ST, Lemey P, et al. Metagenomic sequencing at the epicenter of the Nigeria 2018 Lassa fever outbreak. Science 2019; 363(6422): 74-+.

34. Nachega JB, Sam-Agudu NA, Ogoina D, et al. The surge of mpox in Africa: a call for action. Lancet Glob Health 2024.

