## Supplemental material for "Likely cross-border introductions of MPXV Clade I into the Republic of the Congo from the Democratic Republic of the Congo"

Supplementary material

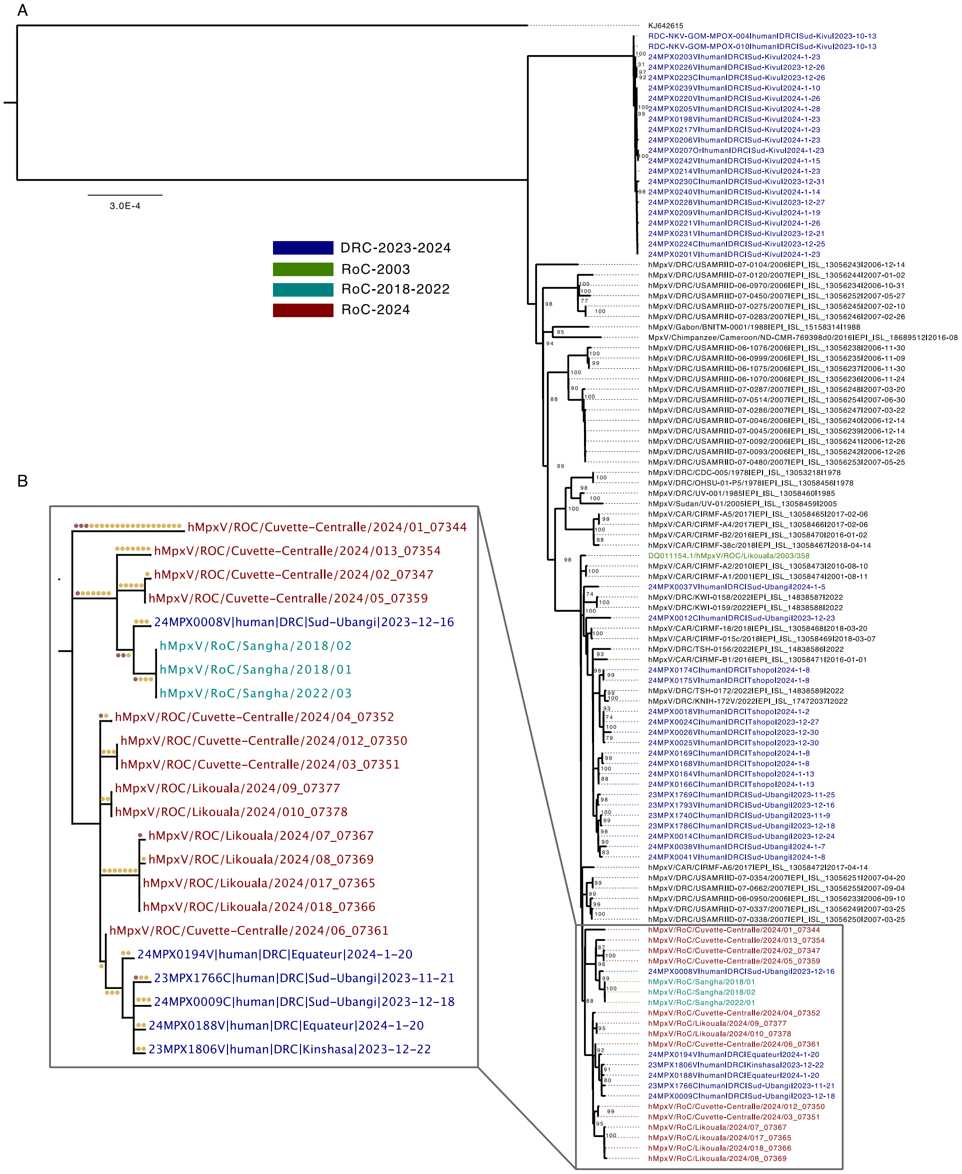


**Supplementary Fig. 1**: Phylogenetic analysis of MPXV full genomes including historic and contemporary Clade I sequence. **A**. Alignments were built with MAFFT (FFT-NS-1 algorithm) [1], with the best model for distance estimates identified with the ModelFinder function [2] as the one with the lowest Bayesian information criterion (BIC). Maximum likelihood phylogenetic tree was constructed using IG-TREE2 [3] and branch support was assessed using both ultrafast bootstrap approximation (ufBoot, 1000 replicates) [4] and SH-like approximate likelihood ratio test (SH-aLRT). The tree was visualized in FigTree (http://tree.bio.ed.ac.uk/software/figtree/) and the phylogenetic tree were rooted using Clade II MPXV sequence KJ642615. MPXV sequences obtained in this study are indicated in red, whereas historic RoC sequences are indicated in green, and contemporary DRC sequences from the current outbreaks are indicated in blue [5]. Node labels represent bootstrap values greater than 69, 1,000 replicates. Scale bar indicates nucleotide substitutions per site. **B**. Ancestral reconstruction performed for each internal node on the phylogeny of the Republic of the Congo cluster and closely related sequences from Democratic Republic of the Congo using IQ-TREE 2, enabling mapping of single nucleotide polymorphisms (SNPs) along branches following the method of O'Toole et al 2024 [6]. SNPs are colored by whether they are consistent with APOBEC3 deamination (red) or not (orange). sub/site, substitutions per site.

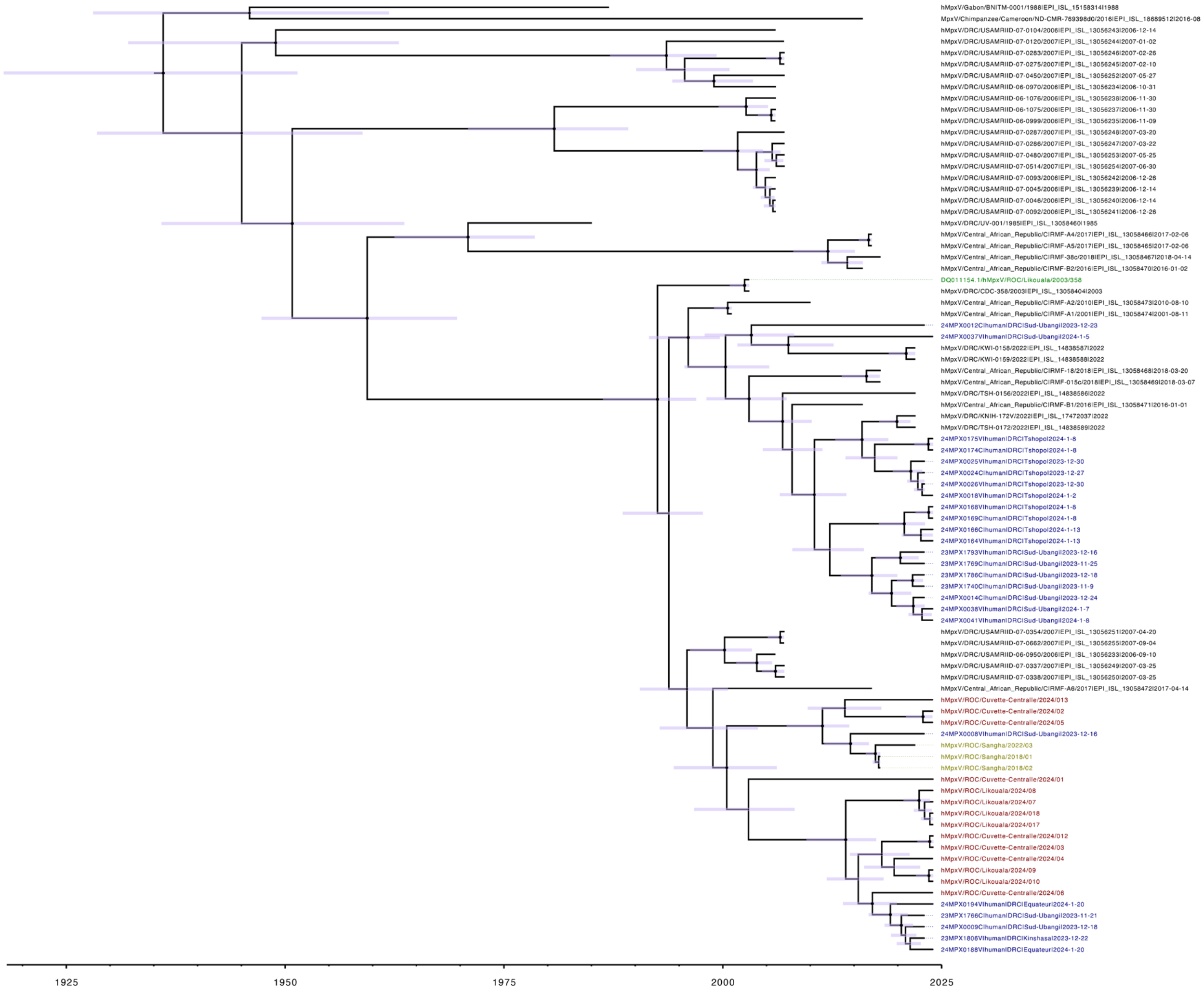


**Supplementary Fig. 2**: **Bayesian phylogenetic inference of MPXV Clade 1a**. Bayesian tree was constructed using a strict clock and constant analysis models. MPXV sequences obtained in this study are indicated in red, whereas historic RoC sequences are indicated in green, and contemporary DRC sequences from the current outbreaks are indicated in blue. Horizontal node bar indicates 95% highest posterior density (HPD) intervals.

References


1. Katoh K, Standley DM. MAFFT multiple sequence alignment software version 7: improvements in performance and usability. Mol Biol Evol 2013;30(4):772-80. (In eng). DOI: 10.1093/molbev/mst010.
2. Kalyaanamoorthy S, Minh BQ, Wong TKF, von Haeseler A, Jermiin LS. ModelFinder: fast model selection for accurate phylogenetic estimates. Nat Methods 2017;14(6):587-589. (In eng). DOI: 10.1038/nmeth.4285.
3. Minh BQ, Schmidt HA, Chernomor O, et al. IQ-TREE 2: New Models and Efficient Methods for Phylogenetic Inference in the Genomic Era. Molecular Biology and Evolution 2020;37(5):1530-1534. DOI: 10.1093/molbev/msaa015.
4. Hoang DT, Chernomor O, von Haeseler A, Minh BQ, Vinh LS. UFBoot2: Improving the Ultrafast Bootstrap Approximation. Mol Biol Evol 2018;35(2):518-522. (In eng). DOI: 10.1093/molbev/msx281.
5. Vakaniaki EH, Kacita C, Kinganda-Lusamaki E, et al. Sustained Human Outbreak of a New MPXV Clade I Lineage in Eastern Democratic Republic of the Congo. medRxiv 2024:2024.04.12.24305195. DOI: 10.1101/2024.04.12.24305195.
6. O’Toole Á, Neher RA, Ndodo N, et al. APOBEC3 deaminase editing in mpox virus as evidence for sustained human transmission since at least 2016. Science 2023;382(6670):595-600. DOI: doi:10.1126/science.adg8116.
